# Effectiveness of Convalescent Plasma for Treatment of COVID-19 Patients

**DOI:** 10.1101/2020.08.02.20166710

**Authors:** Shanshan Chen, Chunya Lu, Ping Li, Lei Wang, Huaqi Wang, Qiankun Yang, Liyinghui Chen, Jianbin Li, Hongwei Ma, Qian Sang, Jing Li, Luyang Xu, Xiangjin Song, Fangfang Li, Yi Zhang, Yi Kang, Lihua Xing, Guojun Zhang

## Abstract

**Background and objective:** The outbreak of COVID-19 has become a global health concern. In this study, we evaluate the effectiveness and safety of convalescent plasma therapy in patients with severe and critically ill COVID-19.

**Methods:** Sixteen COVID-19 patients received transfusion of anti-COVID-19 antibody-positive convalescent plasma. The main outcome was time for viral nucleic acid amplification (NAA) test turning negative. Clinical laboratory parameters were measured at the baseline (d0) before plasma transfusion, and day 1 (d1), day 3 (d3) after transfusion as well.

**Results:** Among the 16 patients, 10 of them had a consistently positive result of viral NAA test before convalescent plasma transfusion. Eight patients (8/10) became negative from day 2 to day 8 after transfusion. Severe patients showed a shorter time for NAA test turning negative after transfusion (mean rank 2.17 vs 5·90, P = 0.036). Two critically ill patients transfused plasma with lower antibody level remained a positive result of NAA test. CRP level demonstrated a decline 1 day after convalescent plasma treatment, compared with the baseline (*P* = 0.017). No adverse events were observed during convalescent plasma transfusion.

**Conclusions:** Viral NAA test of most patients with COVID-19 who received convalescent plasma transfusion turned negative on the 2nd to 8th days after transfusion, and the negative time of severe patients was shorter than that of critically ill patients.

**Trial Registration:** Chinese Clinical Trial Registry; No.: ChiCTR2000030627 URL:http://www.chictr.org

## Introduction

An outbreak of viral pneumonia has been reported in Wuhan City, Hubei Province, P. R. China since December 2019. The disease was subsequently confirmed to be an infectious disease caused by a novel coronavirus infection which is now named as severe acute respiratory syndrome coronavirus 2 (SARS-CoV-2) ^1^, and WHO names the disease induced by SARS-CoV-2 as coronavirus disease 2019 (COVID-19) ^2^. COVID-19 has spread rapidly globally^3-6^ and a total of 82,880 confirmed cases and 4,633 deaths were reported in China before May 3, 2020, with a fatality rate of 5.59%^7^. A retrospective study of 52 critically ill COVID-19 patients in Wuhan Jin Yin-tan Hospital showed that mortality rate of critically ill cases was as high as 51·2% (32/52)^8^. Under the strict and comprehensive measures to control the source of infection and cut off the route of transmission, the number of new cases of COVID-19 in China has decreased significantly. The management of COVID-19 is now focusing on the treatment of severe and critically ill patients.

There are currently no specific antiviral drugs for COVID-19, the clinical effects of some drugs such as lopinavir/ritonavir, chloroquine phosphate, and abidol still need further validation in clinical trials although they have been included in the guideline published by National Health Commission of P. R. China ^9,10^. Convalescent plasma transfusion has been used for clinical treatment of acute viral infectious diseases for a long time, and it has been used in the Spanish influenza pneumonia more than 100 years ago. A meta-analysis shows that patients with Spanish influenza pneumonia transfusion with influenza-convalescent human blood products reduced the risk for death ^11^. Moreover, convalescent plasma has also been used in the treatment of severe acute respiratory syndrome (SARS), Middle East respiratory syndrome (MERS), H1N1 influenza, and Ebola disease^12-15^. The 4th edition of COVID-19 and the later updated clinical diagnosis and treatment scheme by National Health Commission of P. R. China also mentioned that convalescent plasma transfusion can be recommended to treat severe and critically ill patients under the symptomatic treatment, respiratory support, and circulatory support treatment^10,16,17^.

In this study, we analyzed 16 patients diagnosed as COVID-19 who received convalescent plasma in the First Affiliated Hospital of Zhengzhou University and Henan Provincial People’s Hospital to evaluate the safety and effectiveness of convalescent plasma in COVID-19.

## Methods

This case series study was approved by the Ethics Committee of Scientific Research and Clinical Trials of the First Affiliated Hospital of Zhengzhou University and the Henan Provincial People’s Hospital, respectively. The approval number is 2020-KY-060. Patients diagnosed as COVID-19 by the SARS-CoV-2 nucleic acid amplification (NAA) test were given the convalescent plasma treatment if they fulfilled the following criteria: Rapidly progressed, severe or critically ill patient^10^, with a written informed consent given by the patient or next-of-kin. Patients were excluded if they were hypersensitive to plasma or plasma products, were known to have immunoglobulin A deficiency. Patients received a transfusion of convalescent plasma according to the ABO-compatible principle, 200-400 mL convalescent plasma was transfused each time, and could be repeated transfused 2-3 times if necessary. Adverse events were closely observed during transfusion. Conventional treatment including supportive treatment, antiviral treatment, antibacterial treatment, traditional Chinese medicine treatment, and respiratory circulation support treatment, etc. were carried out according to the clinical diagnosis and treatment scheme of COVID-19^10^.

The primary outcome is the time for viral NAA test turning negative (two consecutive negative tests for viral nucleic acid with an interval time not less than 24 hours) after convalescent plasma transfusion. Laboratory parameters at baseline (d0), one day (d1), and three days (d3) after transfusion were recorded, including white blood cell (WBC) counts, neutrophil (NEU) counts, lymphocyte (LYM) counts, C-reactive protein (CRP), procalcitonin (PCT), alanine aminotransferase (ALT), aspartate aminotransferase (AST), Lactate dehydrogenase (LDH), creatine kinase (CK), creatine kinase isoenzyme (CKMB), hypersensitive troponin T (cTnT), and lactic acid (Lac). The ratios of d1/d0, and d3/d0 were calculated to describe the change. Clinical information such as invasive ventilation, application of extracorporeal membrane oxygenation (ECMO), comorbidities were also recorded.

All plasma donors has diagnosed as COVID-19 by NAA test and recovered from infection, continued the quarantine for 2 weeks after discharge. The donors should also meet the following criteria: recovered patients aged 18-55 years old, with the weight more than 50Kg for males and 45Kg for females, no blood-borne diseases, assessed by a clinician to be able to donate plasma, and gave a written informed consent by the donor. The apheresis plasma machine was used to collect the plasma of donors at Henan Red Cross Blood Center (Zhengzhou, China). After the plasma was collected, pathogens were detected and the plasma was stored by the blood station in accordance with the relevant operating procedures. Qualified samples will be sent to the medical institution for detection of plasma antibody level using the method of magnetic particle chemiluminescence.

All data were tested for distribution of normality. Data met the normal distribution were expressed as mean ± standard deviation, and compared using the t test. Data did not meet the normal distribution were expressed as median (minimum, maximum value), and compared using a non-parametric test (Wilcoxon or Mann-Whitney U test). SPSS (version 21.0; SPSS) was used for statistical computation. A *P* value of 0.05 was considered to represent significant difference.

## Results

A total of 16 patients, including 5 female and 11 male patients, received convalescent plasma treatment (**Table 1**). Patients were aged 30-90 years, with an average age of (65 ± 19) years. Among the enrolled patients, 5 of them (31.25%, 5/16) were in a severe illness, and 11 (68.75%, 11/16) were critically illness; 62.50% (10/16) of the patients had comorbidities; the rates of invasive ventilation and use of extracorporeal membrane oxygenation (ECMO) in critically ill patients were 90.91% (10/11) and 45.46% (5/11) respectively. The convalescent plasma transfusion in this study started on the 12th to 36th days of the onset of symptoms, with an average (23 ± 7) days. The plasma transfusion volume ranged from 200 to 1200 mL, with the majority being 200 mL. Upon donation, 200 to 400 mL of convalescent plasma was obtained from 30 donors by apheresis, a cumulative donation of 11.4 L plasma were collected.

**Table 1.** Clinical characteristics of patients received convalescent plasma

| No. | Gender | Age | Severity | Total volume (mL) | Days from symptom onset | Days of use of invasive ventilation | Days of use of non-invasive ventilation | Days of use of ECMO | Comorbidities |
| --- | --- | --- | --- | --- | --- | --- | --- | --- | --- |
| 1 | female | 30 | Critical | 600 | 26 | 10 | N/A | 8 | Gestational diabetes |
| 2 | male | 61 | Critical | 800 | 21 | 4 | N/A | N/A | Coronary heart disease |
| 3 | female | 69 | Critical | 200 | 21 | N/A | N/A | N/A | Diabetes;<br>Postoperative of breast cancer |
| 4 | male | 83 | Critical | 400 | 21 | 12 | N/A | N/A | Coronary heart disease |
| 5 | male | 68 | Critical | 200 | 36 | 1 | N/A | 1 | Chronic bronchitis |
| 6 | male | 73 | Critical | 200 | 29 | 10 | N/A | 2 | N/A |
| 7 | male | 77 | Critical | 1200 | 29 | 9 | N/A | 5 | N/A |
| 8 | male | 62 | Critical | 600 | 22 | 17 | N/A | N/A | Hypertension; cerebral infarction |
| 9 | female | 79 | Severe | 400 | 21 | N/A | N/A | N/A | N/A |
| 10 | male | 40 | Severe | 600 | 33 | N/A | N/A | N/A | N/A |
| 11 | male | 32 | Severe | 600 | 12 | N/A | 1 | N/A | N/A |
| 12 | male | 58 | Critical | 600 | 29 | 11 | N/A | 9 | Hypertension; Coronary heart disease |
| 13 | male | 81 | Severe | 200 | 17 | N/A | N/A | N/A | Chronic bronchitis |
| 14 | female | 87 | Critical | 200 | 18 | 8 | N/A | N/A | Hypertension |
| 15 | female | 46 | Severe | 200 | 14 | N/A | N/A | N/A | N/A |
| 16 | male | 90 | Critical | 200 | 15 | 4 | N/A | N/A | Hypertension; Diabetes; Alzheimer's disease |

This study recorded the time for viral NAA test turning negative after convalescent plasma transfusion (**Table 2**). Viral NAA test of 10 patients were consistently positive before convalescent plasma transfusion. Two critically ill patients remained a positive result of NAA test and were died on the 3rd and 6th day after the transfusion respectively. Eight patients became negative from day 2 to day 8 after transfusion, including 5 critically ill and 3 severe patients. Subgroup analysis demonstrated that severe patients showed a shorter time for NAA test turning negative after transfusion (Mann-Whitney U test, mean rank 2.17 vs 5.90, *P* = 0.036).

**Table 2.** Antibody level and time for viral negation after convalescent plasma transfusion

| No. | Severity | Days for viral negation | Plasma volume (mL) | Median antibody level<br>( AU/mL ) | Accumulated antibody dose<br>(AU) |
| --- | --- | --- | --- | --- | --- |
| 1 | Critical | 3 | 600 | 101.00 | 60,598 |
| 2 | Critical | 8 | 800 | 42.60 | 34,080 |
| 4 | Critical | 5 | 400 | 14.91 | 5,962 |
| 5 | Critical | Remaining Positive | 200 | 16.79 | 3,358 |
| 6 | Critical | Remaining Positive | 200 | 16.79 | 3,358 |
| 7 | Critical | 5 | 600 | 98.63 | 59,176 |
| 8 | Critical | 5 | 600 | 98.63 | 59,176 |
| 9 | Severe | 2 | 400 | 67.20 | 26,880 |
| 10 | Severe | 2 | 600 | 13.58 | 8,146 |
| 11 | Severe | 3 | 600 | 64.10 | 38,458 |

The convalescent plasma antibody level in this study was ranged from 10.93 to 114.7 AU/mL, with an average value of (56.44 ± 39.4) AU/mL. The average plasma antibody level and accumulated dose of antibody before NAA turned negative were calculated (**Table 2**). In 10 severe and critically ill patients, plasma antibody level and accumulated dose showed a similar trend, and the median antibody level of the two consistently positive patients was the lowest (**Figure 1A, 1B**). We then excluded 3 severe patients due to the differences in the time of viral NAA test turning negative between severe and critically ill patients. In 7 critically ill patients, a possible correlation trend was observed between the median plasma antibody level (or accumulated dose) and time of viral NAA test turning negative after convalescent plasma application (**Figure 1C, 1D**). However, this trend was not sufficient to be statistically significant due to the limitation of the number of enrolled patients.

**Figure 1.**
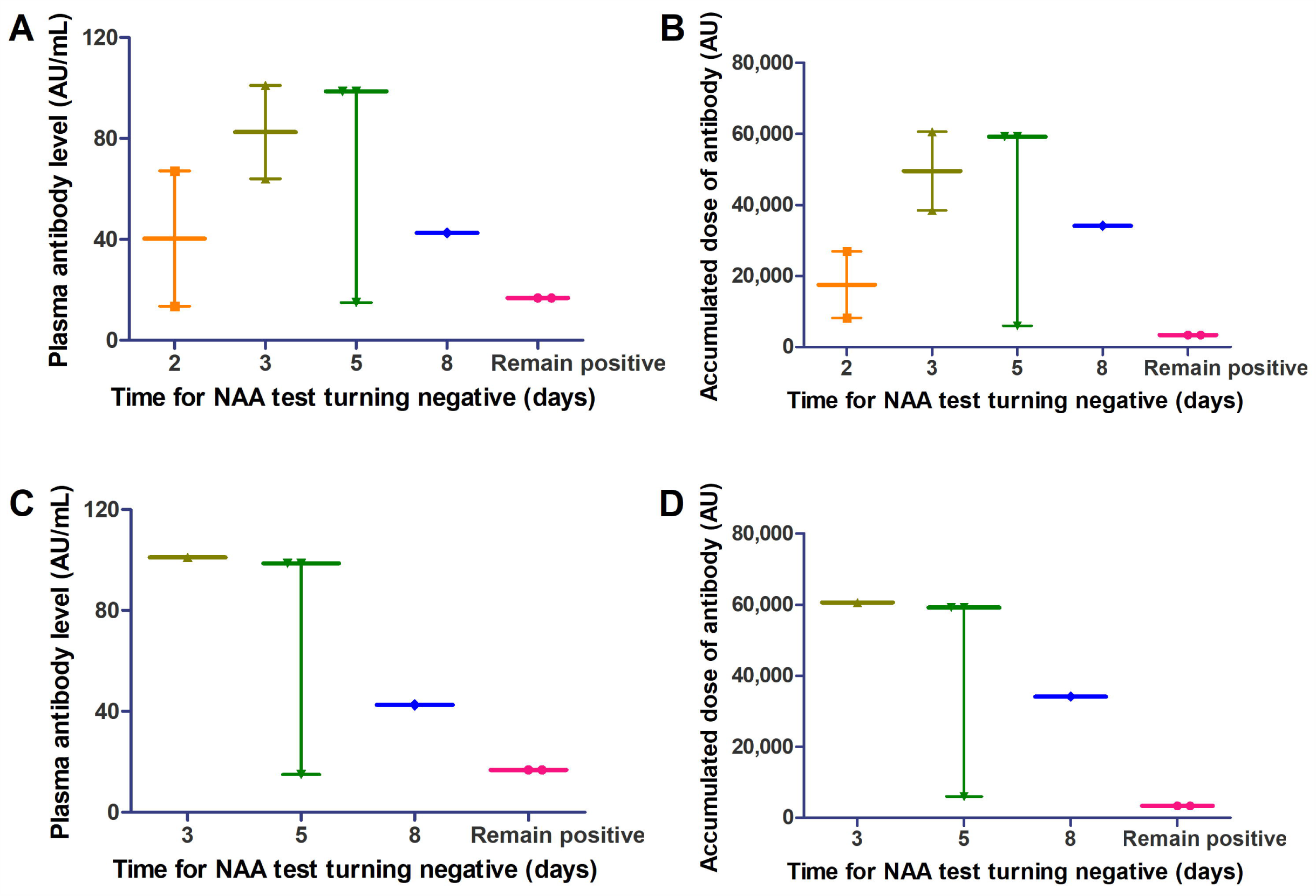
Difference of days for viral NNA test turning negative after treatment of convalescent plasma with different anti-COVID-19 antibody level. (A) and (B) showed the trend in 10 patients in severe and critically illness. (C) and (D) showed trend in 7 critically ill patients. Data were shown as median with range. NAA: nucleic acid amplification.

For laboratory parameters (**Table 3, 4**), CRP level demonstrated a decline 1 day after convalescent plasma treatment with the median ratio of 0.63, compared with the baseline (**Figure 2, 3** *P* = 0.017). WBC (*P*=0.014) and NEU ratio (*P*=0.009) at d1/d0 also showed a decline. No significance was found in the ratio of d3/d0 (Table S1, S2 in appendix; *P*>0.05). No adverse events were found during convalescent plasma transfusion in 16 patients.

**Table 3.** Ratio of d1/d0 of laboratory parameters in patients (Data with normal distribution)

| Parameters | n | Mean | SD | t | P |
| --- | --- | --- | --- | --- | --- |
| WBC | 16 | 0.81 | 0.28 | -2.80 | 0.014* |
| NEU | 16 | 0.75 | 0.33 | -3.00 | 0.009* |
| LYM | 16 | 0.92 | 0.36 | -0.87 | 0.398 |
| AST | 16 | 1.16 | 0.35 | 1.87 | 0.082 |
| CK | 14 | 0.98 | 0.52 | -0.14 | 0.892 |

\* The difference is statistically significant.

**Table 4.** Ratio of d1/d0 of laboratory parameters in patients (Data with non-normal distribution)

| Parameters | n | Median | Minimum | Maximum | P |
| --- | --- | --- | --- | --- | --- |
| CRP | 15 | 0.63 | 0.44 | 1.43 | 0.017* |
| PCT | 14 | 0.86 | 0.49 | 2.72 | 0.683 |
| ALT | 16 | 0.96 | 0.31 | 3.00 | 0.650 |
| LDH | 14 | 0.98 | 0.00 | 1.46 | 0.594 |
| CKMB | 15 | 0.84 | 0.00 | 4.59 | 1.000 |
| cTnT | 10 | 0.94 | 0.00 | 8.50 | 0.878 |
| Lac | 8 | 0.95 | 0.64 | 2.11 | 0.674 |

\* The difference is statistically significant.

**Figure 2.**
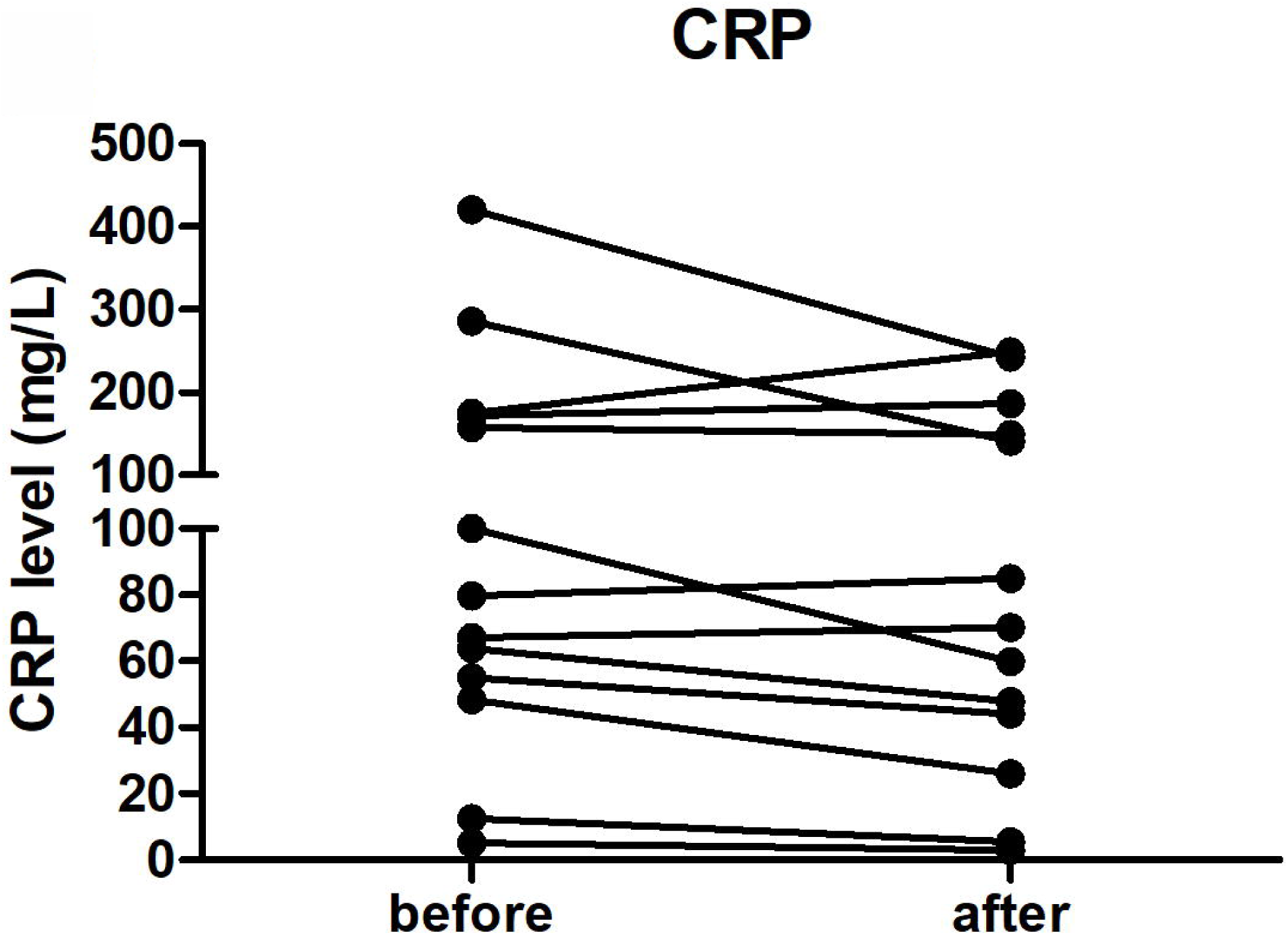
Comparison of CRP level between time of before and one day after convalescent plasma. CRP level showed a decline trend one day after convalescent plasma.

**Figure 3.**
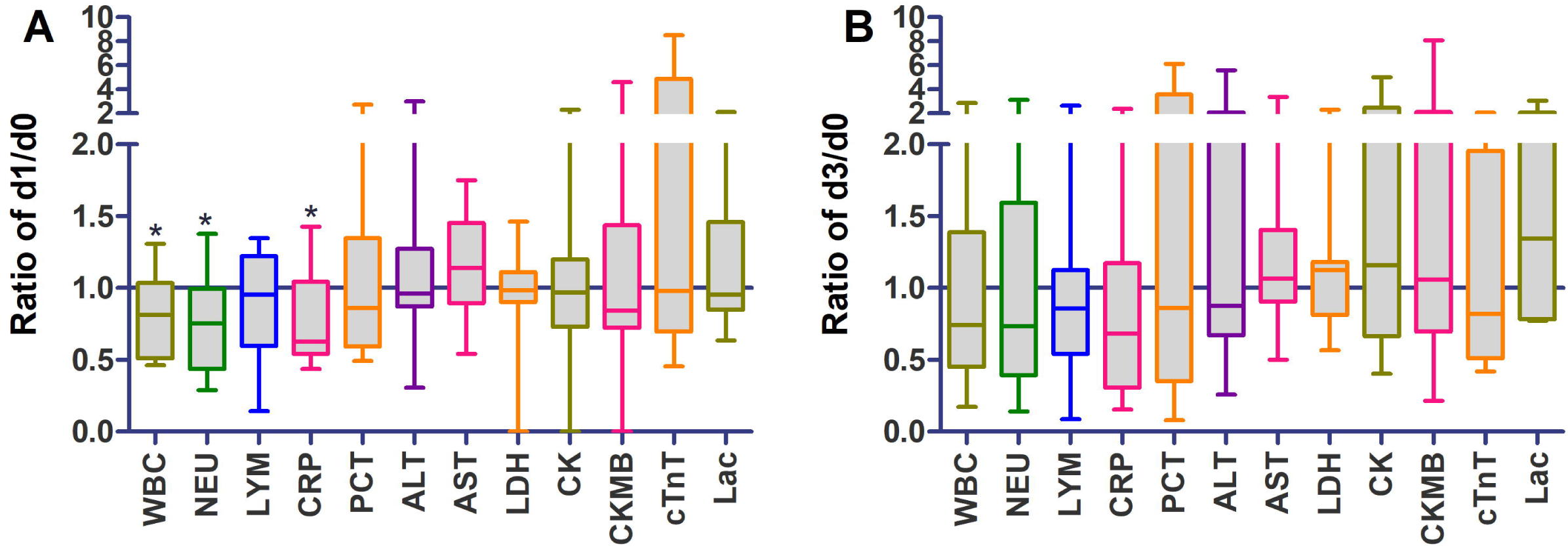
Change of clinical laboratory parameters after convalescent plasma transfusion. (A) Ratio of one day after convalescent plasma transfusion (d1) with baseline (day 0, d0). (B) Ratio of three day after convalescent plasma transfusion (d3) with baseline (day 0, d0). Data were expressed as box and whiskers (minimum to maximum). * represents a significant difference of *P*<0.05.

## Discussion

This study analyzed and summarized the basic characteristics and laboratory parameters of patients with convalescent plasma transfusion in provincial medical institutions in Henan province, China. More than 1,000 COVID-19 patients have been diagnosed in the area. Not only in China, COVID-19 epidemic has become an international public concern and affected more than 90,000 people worldwide^18,19^. We preliminary evaluated some of the effectiveness and safety of convalescent plasma treatment. Shortening viral carrying time is particularly important for the treatment and control of COVID-19. In our study, time for viral NAA test negation is an important observational parameter for evaluating the effectiveness of convalescent plasma in this study. We found that the time of viral negativity was 2-8 days after transfusion for most enrolled patients (8/10), in addition, this time was even shorter in severe patients than in critically ill patients.

As a sensitive indicator of inflammation, CRP is widely used in the evaluation of various infectious diseases alone or in combination with other indicators^20,21^. Increased CRP level was observed in 93.75% (15/16) of the patients in the baseline, and the average CRP level of patients was observed to drop to about 0.63 of the baseline level, suggesting convalescent plasma transfusion may play a role in reducing the body’s inflammatory response rapidly. In addition, in this study, a decrease in the counts of WBC and neutrophils was observed one day after plasma transfusion. The original data showed the WBC count of 12 patients (75%) were within the normal range in the baseline, and WBC counts of most patients fluctuated within the normal range. Therefore, we believe that the changes of WBC and neutrophils we observed are not clinically significant in evaluating the effectiveness in convalescent plasma.

It is worth noting that 2 patients (2/10) remained positive in virus NAA test after convalescent plasma transfusion, and the disease progressed rapidly and finally caused death in a short period of time, indicating the function of convalescent plasma in some patients may be very limited. In order to make sure the safety of plasma samples, all plasma donors has recovered from infection and continued the quarantine for 2 weeks after discharge. As fewer new cases with severe or critically illness in our province were reported in our province, early use of convalescent plasma is limited, most of the patients received convalescent plasma treatment in this study were critically ill patients who had a history of ventilator and ECMO applications, which may attenuate the effects of convalescent plasma. During the 2009 H1N1 influenza epidemic, 20 patients has enrolled in a convalescent plasma clinical trial reported by Hung et al. In their study, the plasma neutralizing antibody titer (NAT) was greater than 1: 160, and the enrolled patients were admitted to the ICU within 7 days of the onset of the disease, convalescent plasma transfusion was performed on the 2nd day on average. Hung et al. concluded that convalescent plasma therapy can reduce mortality in patients with H1N1 infection (20.0% vs 54.8%; *P*=0.01)^22^. In another multicenter, double-blind, randomized controlled trial, 18 patients were treated with hyperimmune IV immunoglobulin (H-IVIG, product from convalescent plasma) within 5 days of the onset of symptoms with H1N1 infection, which could also reduce the viral load and death of patients rate^23^. Therefore, early use of convalescent plasma therapy is recommend in COVID-19 patients. However, there is also a study for Ebola disease showed that two consecutive transfusions of convalescent plasma for a dose of 200-250 mL within 2 days of onset did not improve disease survival^13^.

In addition to the early application of convalescent plasma therapy, concentration of antibody in convalescent plasma may be another factor closely associated with effectiveness. Two patients died with a persistently positive viral NAA test after plasma transfusion in the study, the plasma (200 mL for each) they transfused came from a same donor with an low antibody concentration of 16.79 AU/mL. Our small sample study found that there may be a correlation between the plasma antibody concentration and the time for viral turning negative in critically ill patients, patients transfused plasma with a higher antibody level may have a shorter time for viral test negation. In the above-mentioned paper, convalescent plasma treatment showed a negative conclusion in Ebola disease, we noticed the researchers did not detect and screen the antibody level in the plasma^13^. Brown J F et al. proposed that convalescent plasma with higher anti-Ebola antibody levels may have a greater impact on the Ebola virus load in patients with Ebola disease^24^. Therefore, the level of specific antibody in convalescent plasma may be an important determinant of the efficacy of plasma therapy. Testing and screening plasma with high level of antibody, and applying in severe or critically ill patients with early stage are expected to improve the efficacy of convalescent plasma.

## Conclusions

Viral NAA test of most patients with COVID-19 who transfused convalescent plasma turned negative on the 2nd to 8th days after transfusion, and the negative time of severe patients was shorter than that of critically ill patients. More clinical trials with larger participants are needed to validate the effectiveness of convalescent plasma.

## CRediT author statement

**Shanshan Chen:** Data curation, Investigation, Project administration, Writing-review & editing. **Chunya Lu:** Methodology, Software, Writing-original draft. **Ping Li:** Data curation, Project administration, Software, Validation, Writing-review & editing. **Lei Wang:** Conceptualization, Resources, Software, Supervision, Writing-review & editing. **Hua-qi Wang:** Conceptualization, Data curation, Project administration, Writing-review & editing. **Qiankun Yang:** Data curation, Resources, Writing-review & editing. **Liyinghui Chen:** Data curation, Resources, Software, Writing-review & editing. **Jianbin Li:** Conceptualization, Resources, Supervision, Writing-review & editing. **Hongwei Ma:** Data curation, Resources, Validation, Writing-review & editing. **Qian Sang:** Investigation, Supervision, Validation, Writing-review & editing. **Jing Li:** Data curation, Methodology, Validation, Writing-review & editing. **Luyang Xu:** Data curation, Project administration, Resources, Writing-original draft. **Xiangjin Song:** Data curation, Investigation, Validation, Visualization, Writing-review & editing. **Fangfang Li:** Data curation, Investigation, Resources, Visualization, Writing-review & editing. **Yi Zhang:** Conceptualization, Data curation, Project administration, Resources, Supervision, Writing-review & editing. **Yi Kang:** Conceptualization, Project administration, Resources, Writing-review & editing. **Lihua Xing:** Conceptualization, Data curation, Resources, Supervision, Writing-review & editing. **Guojun Zhang:** Conceptualization, Funding acquisition, Project administration, Supervision, Writing-review & editing.

## Data Availability

The data used to support the findings of this study are available from the corresponding author upon request.

## Acknowledgements

This work was funded by Science and Technology Department of Henan Province. (Emergency research project for prevention and control of COVID-19, No.2011003 11000).

## Declarations of interest

None.

## Notes

### Competing Interest Statement

The authors have declared no competing interest.

### Clinical Trial

ChiCTR2000030627

### Author Declarations

This case series study was approved by the Ethics Committee of Scientific Research and Clinical Trials of the First Affiliated Hospital of Zhengzhou University and the Henan Provincial People's Hospital, respectively. The approval number is 2020-KY-060.

